# Medical Capacity Shortages Facilitated the Rapid Dissemination of COVID-19 in Wuhan, New York State, and Italy

**DOI:** 10.1101/2020.11.05.20226530

**Authors:** Yuehao Xu, Cheng Zhang, Lixian Qian

## Abstract

During the coronavirus disease 2019 (COVID-19) outbreak, every public health system faced the potential challenge of medical capacity shortages. Infections without timely diagnosis or treatment may facilitate the stealth transmission and spread of the virus. Using infection and medical capacity information reported in Wuhan in China, New York State in the United States, and Italy, we developed a dynamic susceptible–exposed–infected–recovered (SEIR) model to estimate the impact of medical capacity shortages during the COVID-19 outbreak at the city, state, and country levels. After accounting for the effects of travel restrictions and control measures, we find that the number of infections in Wuhan could have been 39% lower than the actual number if the medical capacity were doubled in this city. Similarly, we find the less shortages in medical capacity in both New York state and Italy, the faster decline in the daily infection numbers and the fewer deaths. This study provides a method for estimating potential shortages and explains how they may dynamically facilitate disease spreading during future pandemics such as COVID-19.

## 1. Introduction

The global spread of coronavirus disease 2019 (COVID-19) has overwhelmed health systems early this year [1] and many European countries are now facing the second wave of the pandemic [2]. In the first wave, many public health systems faced the severe challenge of medical capacity shortages. For instance, unexpected critical shortages of hospital beds and medical staff occurred in many countries during the COVID-19 outbreak. Such limited medical capacity imposes an upper bound on the number of patients of being promptly diagnosed and properly treated. Infections without timely diagnosis or treatment may further facilitate the stealth transmission and spread of the virus.

Today, despite the importance of medical capacity, it is unclear to public health decision makers how the abovementioned shortages would exacerbate the spreading of diseases during pandemics such as the COVID-19. On the basis of existing research on the transmission pattern of the epidemic [e.g., 3, 4-7], we developed a dynamic susceptible–exposed–infected–recovered (SEIR)-based inference framework to estimate the impact of medical capacity shortage on the spread of COVID-19 during the pandemic. We used the data in the first wave of COVID-19 pandemic in Wuhan city, New York State and Italy to calibrate the model. By doing so, we demonstrate that our model can be used by the public health decision makers to better prepare medical resources for the outbreak of second wave of COVID-19 as well as other epidemics in future.

Generally, we denoted *S*(*t*),*E*(*t*), *I*(*t*), and *R*(*t*) as the susceptible, exposed, infected, and recovered numbers at time *t*, respectively. In contrast to the assumption that all infected patients were hospitalized immediately, health systems were rapidly overwhelmed at the beginning of the COVID-19 outbreak [1], with the medical capacity in terms of the hospitalized rate decreasing sharply after a very short period. Subsequently, overwhelmed health systems constantly operated at a low level and were unable to recover rapidly because the incremental rate of confirmed infections exceeded the recovery rate. In such a situation, health systems could not promptly diagnose and treat infected patients. After the control measures and treatments took effect, the recovery rate of patients would exceed the newly infected rate and then the hospitalized rate recovers gradually until it reaches its normal ceiling. However, the recovery of medical capacity was slow, because critically ill patients, particularly elderly ones, take a substantially longer time to recover than do other patients [8] and an increasing proportion of critically ill patients receive hospital care in the later stages of disease progression.

Infectious disease usually spreads through social contagion with a networked effect [9]. Thus, the transmission of the virus grows exponentially at the beginning and then slows down when the pandemic is gradually under control. Given such pattern, the hospitalized rate would increase in an S-shaped sigmoid curve until *T*_1_, when the shortage in medical capacity would be resolved with all medical and nonmedical interventions exerting their maximum effect. After *T*_1_, the rate would remain constant. Thus, we modeled the hospitalization rate *fraction*(*t*) as a piecewise function: it follows a Gompertz function [10] until *T*_1_, and then remains constant thereafter:

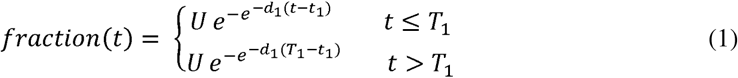

where *U* is the saturation level of hospitalization rate, *d*_1_ is the decay parameter to be fitted in the model and *t*_1_ is the inflection point of the S-shaped curve where the rate equals 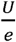, which may be different (i.e., earlier or later) from the day the measures took effect. *T*_1_ is the time point that the fraction does not change any more. The Gompertz function is characterized by its asymmetric inflection at about 36.8% (=1/*e*) of the saturation level, which can reflect the difficulties in recovering the hospitalization rate after the medical capacity shortage occurred in the early stage. In addition, without general loss, *U* is the extrinsic factor that may also affect the overall hospitalized rate. For instance, the United States and early-stage Italy implemented a home-staying policy and only confirmed infections with severe symptoms were hospitalized. In regions where health care is intensively provided at designated locations, such as South Korea and China, all confirmed infections must be treated in hospitals or special facilities and should not be home-isolated. Countries such as Singapore have even employed preventative swab tests to identify more infected patients and isolate them.

Furthermore, various non-pharmaceutical interventions (NPIs), such as maintaining social distancing, promoting good hygiene practices, and enhancing supply chains, can slow transmission over time [11-13]. Accordingly, we built the following piecewise transmission parameter function *β*(*t*) in another Gompertz function form to reflect the transmission dynamics

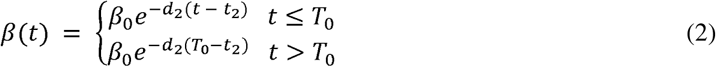

where *d*_2_ is the decay parameter of the transmission rate that would be fitted in the model. The NPI measures are introduced on day *t*_2_ and reach their ceiling point on day *T*_0_. Therefore, the transmission parameter *β*(*t*) follows a monotonic decreasing pattern until *T*_0_ and then remains constant thereafter.

Therefore, we developed a time-decaying SEIR model, which consists of the following four differential equations:

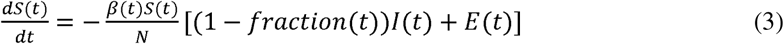

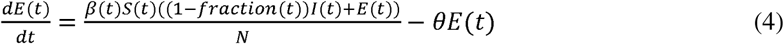

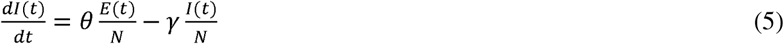

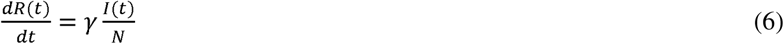

where *β*(*t*) denotes the piecewise transmission rate at day *t* as specified in eq.(2), and (1− *fraction*(*t*))*I*(*t*) stands for the number of infected cases that have not been hospitalized because of medical capacity shortages. *N* denotes the population in a specific region (i.e., a city, state, or country), *θ* denotes the incubation rate (i.e., the rate of conversion from exposed to infected per day), and *γ* denotes the recovery rate per day.

## 2. Data and Model Parameters

We first apply the proposed model in the epidemic case of Wuhan city (the provincial capital of Hubei Province) of China during the outbreak of COVID-19 early this year, and then extend the analysis to the New York State and Italy in their first wave of epidemics in March to May 2020. Specifically, we used observations of reported infection and medical capacity information in China to estimate the impact of medical capacity on the spread of COVID-19. The data set included the daily cumulative numbers of infections in Wuhan and other regions of China outside of Hubei Province between January 14 and March 2, 2020.

On the basis of available reports of COVID-19 treatment [14], we initially assumed *γ* and *θ* in the SEIR model to be 0.08 and 0.25, respectively, which mean an average recovery period of 12.5 days and incubation period of 4 days. Medical capacity was proxied by daily bed numbers reported by the Wuhan Health Commission from February 2 to February 25, 2020.

We used the Nelder–Mead method [15] to search our solution for minimizing loss. This method can address multidimensional unconstrained optimization problems that do not have derivatives, which assures its robustness to some degree. We defined the loss function as the sum of squared errors by day:

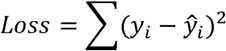

where *y*_*i*_ and *ŷ*_*i*_ denote the real and predicted numbers of infected cases, respectively.

In the case of COVID-19 transmission in China, we denoted January 1, 2020, as the starting point of our model (*t* = 1). The end point for the hospitalization fraction recovery (i.e., *T*_1_) was fixed at *t* = 60 (i.e., March 1, 2020) because almost all identifiable patients had been hospitalized by then. In late January and early February 2020, Wuhan faced a more severe threat from medical capacity shortages than other regions in China. Specifically, before Hubei Province announced its Level 1 emergency response on January 24, the highest level for a public health emergency, Wuhan only had 7,500 hospital beds available to treat infected patients, which was well below the peak number of 35,000 COVID-19 patients in this city. In comparison, because of efficient mobilization, strict quarantine measures and a policy of accepting all patients, the epidemic outside Hubei Province was controlled to a relatively mild level with the sufficient capacity. According to epidemic statistics, there were about 10,000 patients at the peak and 32,717 total infections out of the population of 1.33 billion in non-Hubei regions in China by April 2, 2020, far below the 600,000 beds available outside of Hubei Province^1^. Fig. 1 summarizes the bed shortage situation in Wuhan in February 2020. It shows that the hospitals in Wuhan increased the number of available beds from 7,500 in late January to 25,000 in late February. During the process, the bed occupation rate decreased from 100% to 80%. This dramatic drop occurred between *t* = 45 (February 14, 2020) and *t* = 55 (February 24, 2020).

**Fig. 1.**
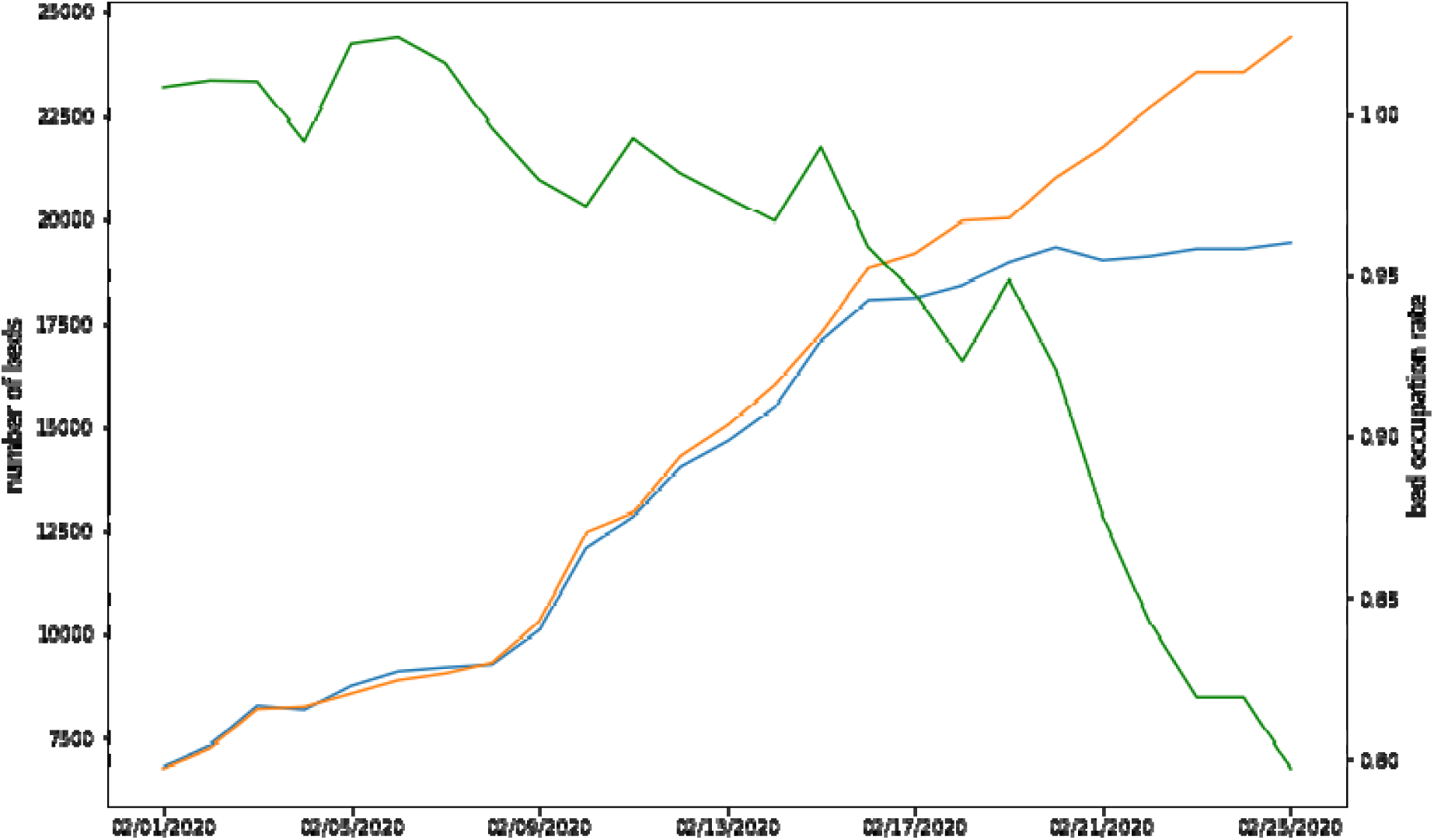
Trend of the capacity shortage in Wuhan. Note: The blue, orange, and green lines denote the daily number of used beds, total number of beds, and occupancy ratio of beds, respectively. The bed occupancy ratio could exceed one in cases of temporary expansion, such as the placement of beds in hallways.

Because there was clearly no bed shortage outside of Hubei Province, we fixed *d*_2_ in eq. (1) at 10 for regions outside of the province; thus, the hospitalized rate in non-Hubei regions was near one, meaning that all infected patients in these places could be hospitalized immediately and none would further infect others in the system. The estimated parameters were *β*_0_,*d*_1_,and *d*_2_ in Wuhan and *β*_0_, *d*_1_,and *R*_*i*_ outside of Hubei Province.

We also use *t*_0_ to denote the starting point of our quantitative analysis on a specific region. When estimating the model, we set the starting point at *t*_0_ = 14 (i.e., January 14, 2020) for the following three reasons. First, it enabled us to capture the most critical period for the spread of COVID-19 in China. Second, the number of infected patients being hospitalized on that day exceeded 250 [16], which was the full early-stage capacity in Wuhan. Third, this date was only 10 days before Chinese New Year in 2020, which means it was the Spring Festival travel season (*chunyun*) for hundreds of millions of passengers traveling from coastal cities (such as Beijing, Shanghai, and Shenzhen) and railway interchange cities (such as Wuhan) to less developed regions [3].

Moreover, the hospitalized rate, defined in eq. (1), increased in an S-shaped curve until *T*_1_ when the hospitalization fraction reached it ceiling (i.e., March 1 in the case of Wuhan). According to eq. (2), the transmission rate decreased negative exponentially until *T*_0_ and remained constant subsequently. Specifically, the turning point *T*_0_ for the transmission rate was set as day *t* = 48 (i.e., February 17, 2020) because it was the time point by which all mobile field hospitals had been built in Wuhan with all measures implemented. Furthermore, we selected the date of NPI implementation *t*_2_ = 24 (i.e., January 24) for Wuhan and *t*_2_ = 28 (i.e., January 28) for the rest of China, because January 24 was when Wuhan went into lockdown and January 28 was the approximate date the whole country entered lockdown. For the inflection point on the shortage of medical capabilities occurred, we selected *t*_1_ = 40 (i.e., February 9) for Wuhan when the hospitalized rate was approximately 0.37 (i.e., 1/*e*). This is because it was the period of the most rapid increase in medical capacity in Wuhan with the construction of mobile field hospitals and quarantine zones.

## 3. Results

Model estimation confirmed the situation in Wuhan: Fig.2 shows that the hospitalized rate for infected patients increased from 0.005 on February 5 to 0.51 on February 10 and to 0.83 on February 13 before it finally exceeded 0.99 after February 21 (see Fig. 2). In contrast to Wuhan, other regions outside of Hubei Province did not face challenges of medical capacity shortage, because the abundant number of beds in hospitals always exceeded the number of locally confirmed cases.

**Fig. 2.**
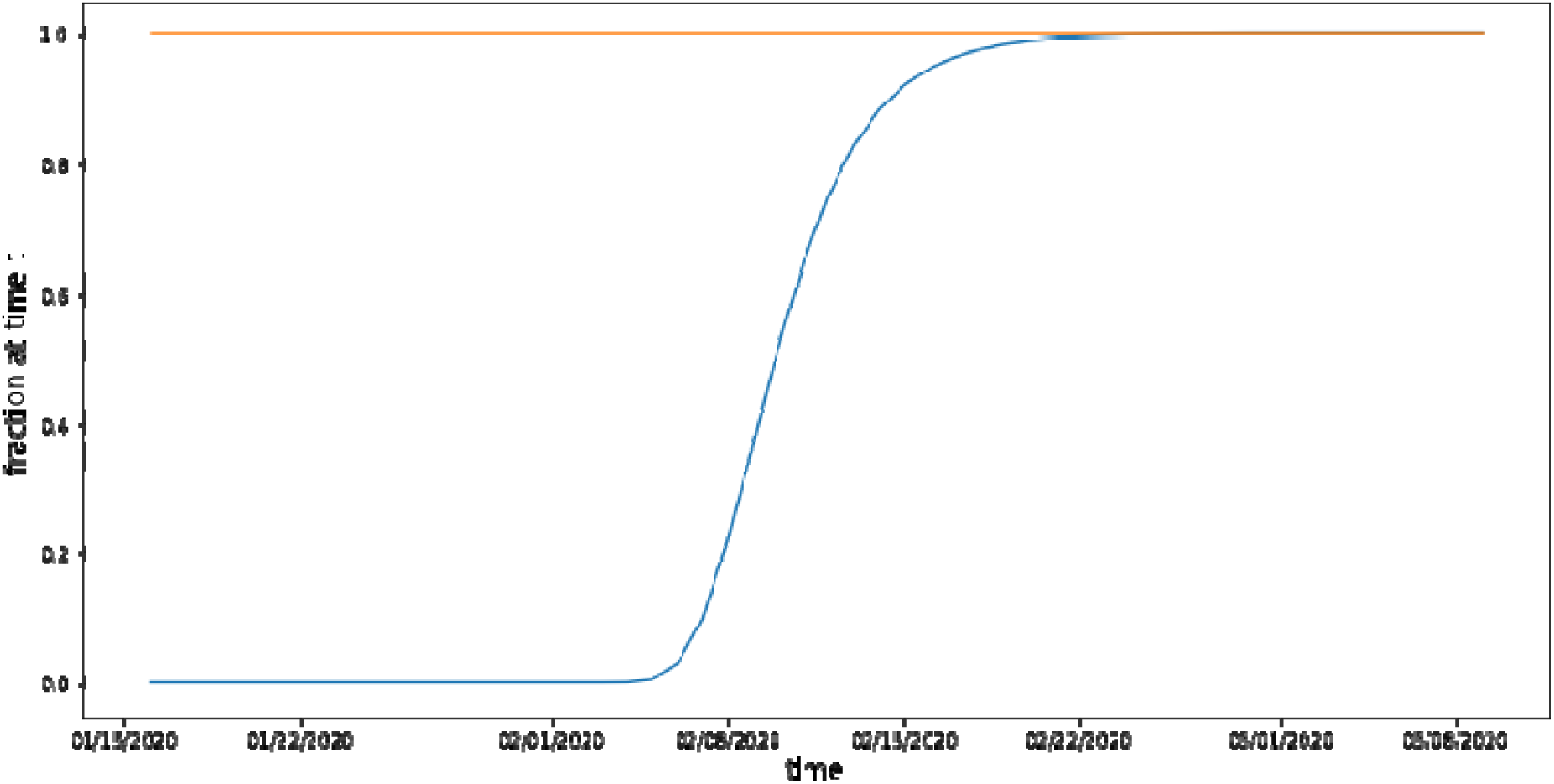
Rate of hospitalization in Wuhan and the rest of China. Note: The blue and orange lines denote the hospitalized rate in Wuhan and outside of Hubei province, respectively. The capacity outside of Hubei Province is always more than the infection number; thus, the rate is one.

The decay in transmission rate, *β*(*t*), also applies to both Wuhan and all other regions in China. Fig. 3 compares the transmission rate in Wuhan (mean value = 0.238, std = 0.08) with that in other regions (mean value = 0.230, std = 0.081). With the same control measures implemented in all regions of China, the difference in transmission rates between Wuhan and other regions reveals the potential effect of reaching the capacity limit on transmission.

**Fig. 3.**
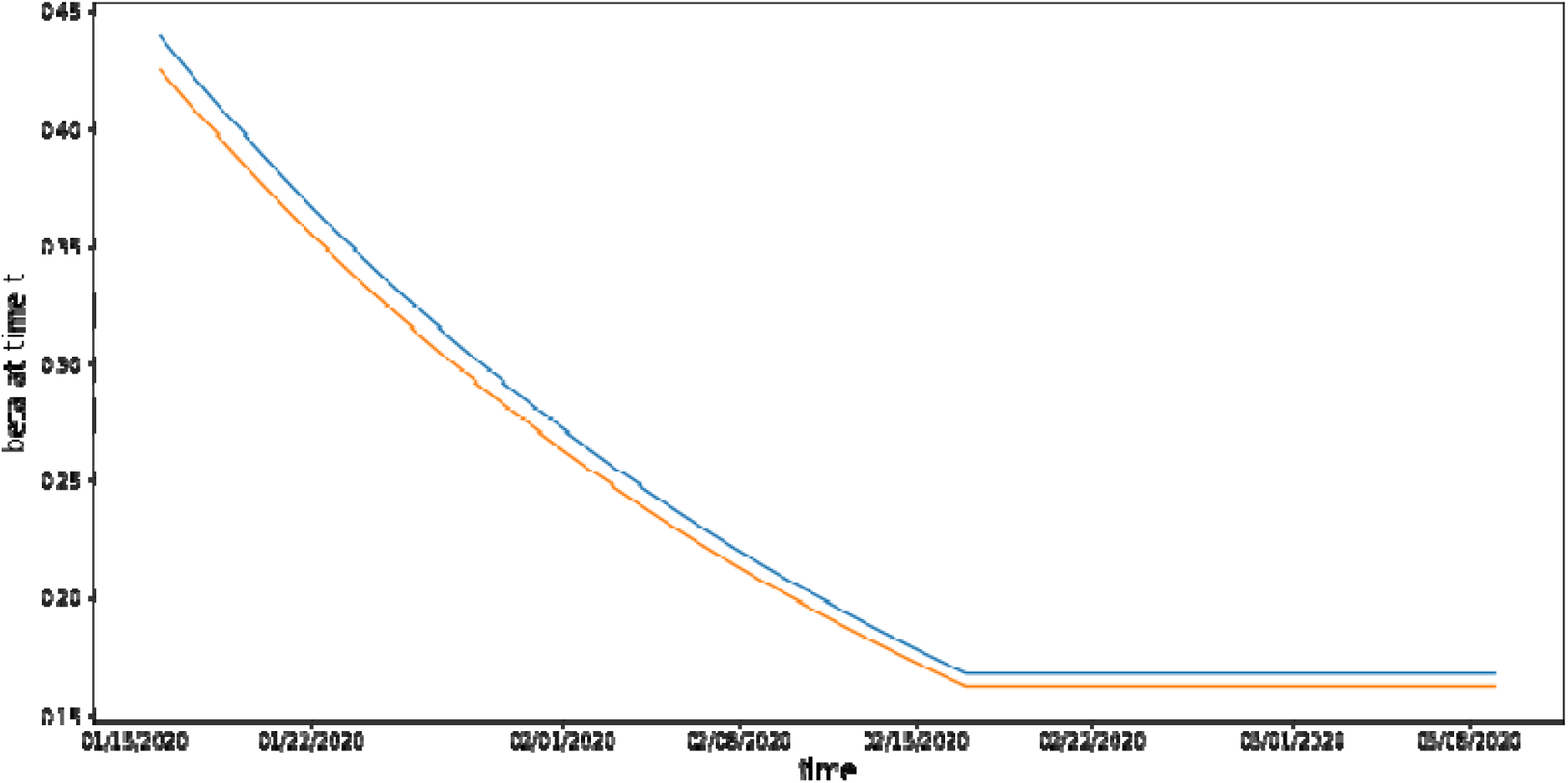
Transmission rate in Wuhan and the rest of China. Note: The blue and orange lines denote the transmission rate in Wuhan and outside of Hubei Province, respectively.

Fig. 4 illustrates the model estimation results for Wuhan. Overall, the model’s explanatory power *R*^2^ was 0.984, with an estimated rate decay rate of *d*_1_ = 0.417 and beta decay rate of *d*_2_ = 0.030 in Wuhan, suggesting that the average treated fraction^2^ increased from 0.10 on February 7 to 0.92 on February 15, 2020. Thus, the model suggested that the medical capacity shortage caused a 32-day delay in timely hospitalization compared with the starting point of January 14. Furthermore, the cumulative number of unhospitalized day-person between January 16 and February 15 was 106,516. If the 12-day recovery period is considered, which means a recovery rate of 8.33% of patients per day, then the total number of unhospitalized people was 8,873 in that month-long period until February 15, 2020.

**Fig. 4.**
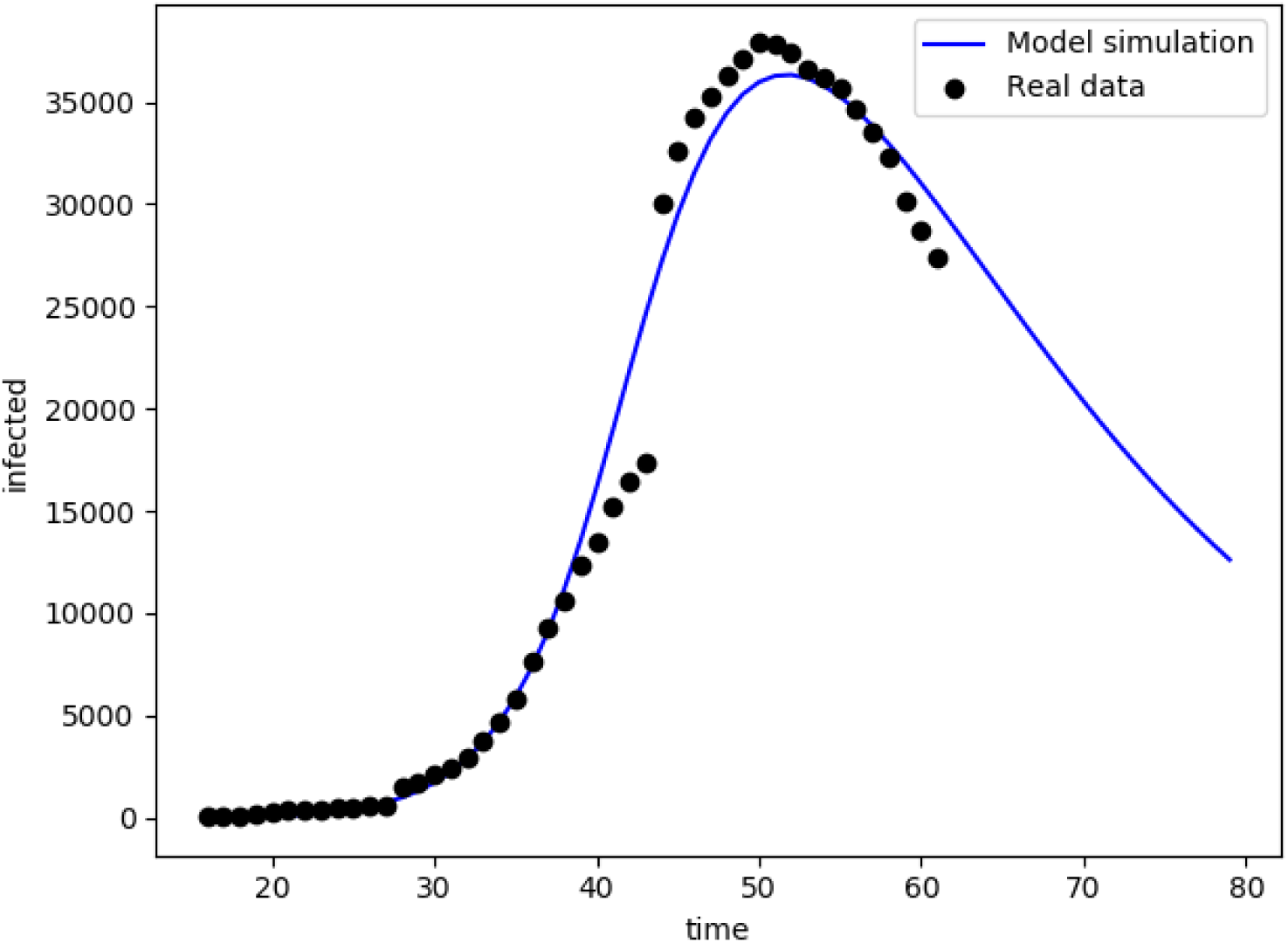
Estimation performance of the model in Wuhan case.

## 4. Sensitivity Analysis in Wuhan Case

We conducted two sensitivity analyses in the context of Wuhan, with respect to the hospitalized rate and transmission rate.

First, we simulated the number of infections in Wuhan due to the different levels of medical capacities. Specifically, we varied the hospitalized rate to be 1.5 times, 1.75 times, or twice of the original rate estimated by our model presented in Fig. 2. As illustrated in Fig. 5, the sensitivity analysis revealed that a 50% increase in medical capacity compared with the original rate would lead to a 27% decrease in the peak number of patients, which dropped from over 36,327 to 28,505 in Wuhan. In addition, 75% and 100% increases in the hospitalized rate would lead to 34% and 39% fewer patients in peak number, respectively.

**Fig. 5.**
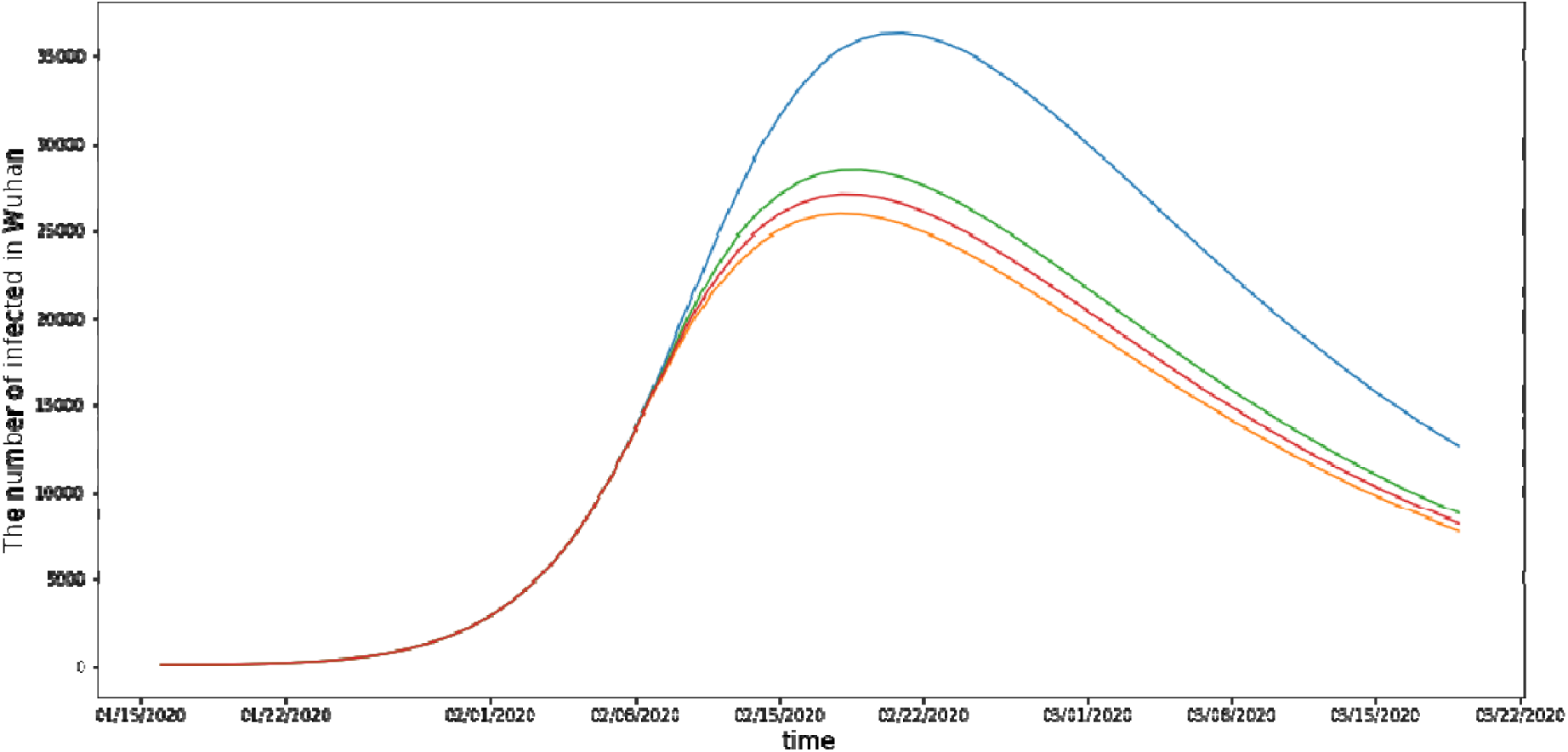
Sensitivity analysis of the hospitalized rate in Wuhan. Note: The blue, green, red, and orange lines denote the predicted number of infected patients in Wuhan with the original hospitalized rate, a 50% increase in the hospitalized rate, a 75% increase in the hospitalized rate, and a 100% increase in the hospitalized rate.

We also conducted a sensitivity analysis on how the transmission rate may worsen or attenuate the effect of reaching the capacity limit. We varied the speed at which the original transmission rate decreased by assuming 0.5, 0.75, 1.25, 1.5, 1.75 times, or twice of the original *d*_1_ value. Fig. 6 illustrate the results. First, we find that, with the given capacity limit in Wuhan on February 19, a 50% slower decay rate d_1_ in transmission would lead to a 175% increase in the number of infected patients and a 25% decrease in the decay rate (i.e., 75% of the original level) would lead to a 26% increase in the number of infected patients. The further sensitivity analysis reveals that a 25%, 50%, 75%, or 100% increase in decay rate d_1_ would lead to 47%, 59%, 66%, or 70% fewer infected patients, respectively. By contrast, Fig. 7 demonstrates that the change in decay rate in regions outside of Hubei Province, where there were no shortages in medical capacity, had a smaller impact on the epidemic compared with in Wuhan. Specifically, a 50% or 25% smaller decay rate *d*_1_ would lead to increases of 34.19% or 9.63% in the number of infected patients respectively, while increases of 25%, 50%, 75%, and 100% in the decay rate would lead to 9.00%, 15.24%, 18.98%, and 21.75% fewer patients respectively. These results indicate the influence of hospitalization capacity on the transmission rate when all control measures are equal; that is, a lower capacity would intensify the influence of different transmission-decay levels, thus influencing the epidemic progression.

**Fig. 6.**
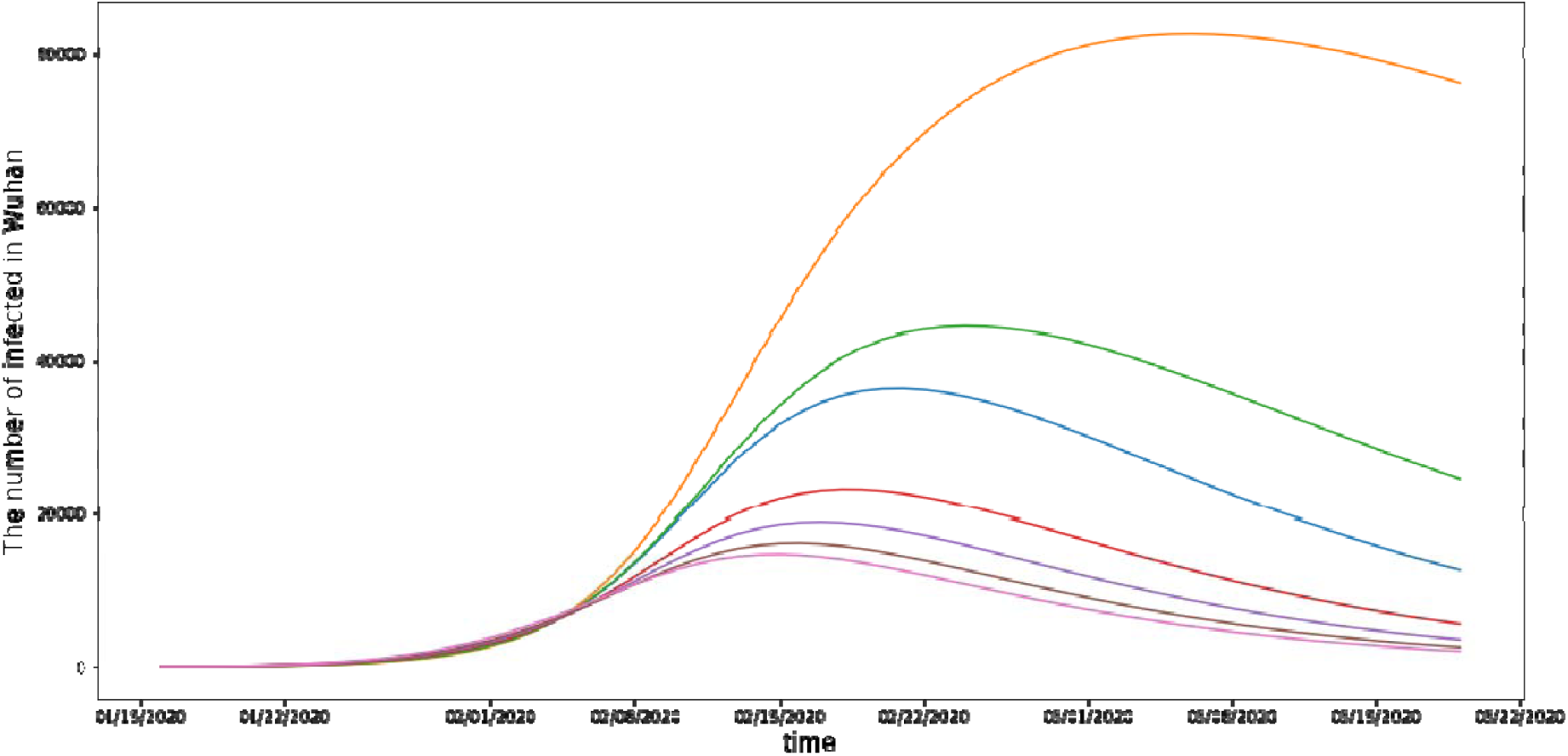
Sensitivity analysis of the transmission rate in Wuhan. Note: The blue, orange, green, red, purple, brown, and pink lines denote the number of infected patients predicted in Wuhan with the original, a 50% lower, a 25% lower, a 25% higher, a 50% higher, a 75% higher, and a 100% higher, respectively.

**Fig. 7.**
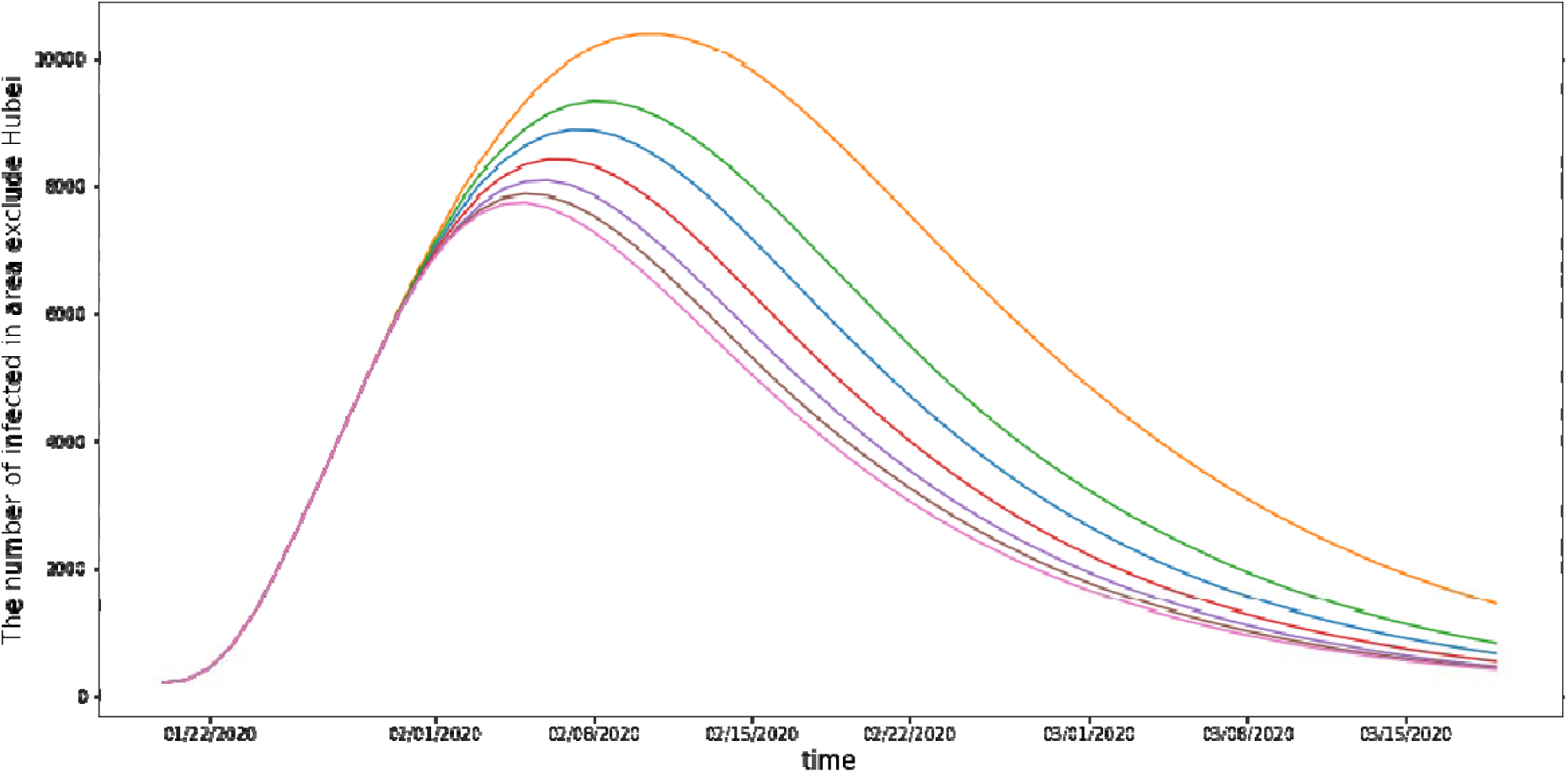
Sensitivity analysis of transmission rate in non-Hubei regions. Note: The blue, orange, green, red, purple, brown, and pink lines denote the predicted number of infected patients in non-Hubei regions with the original, a 50% lower, a 25% lower, a 25% higher, a 50% higher, a 75% higher, and a 100% higher, respectively.

Capacity has also affected the total number of deaths throughout the epidemic. From data provided by the Centers for Disease Control in Wuhan, we estimated the death rate in Wuhan to be 0.05. Moreover, we found that with 1.5 times, 1.75 times, and twice the original rate, the number of deaths would decrease from 4430 to 3474, 3309, and 3179, respectively (see Fig. 8).

**Fig. 8.**
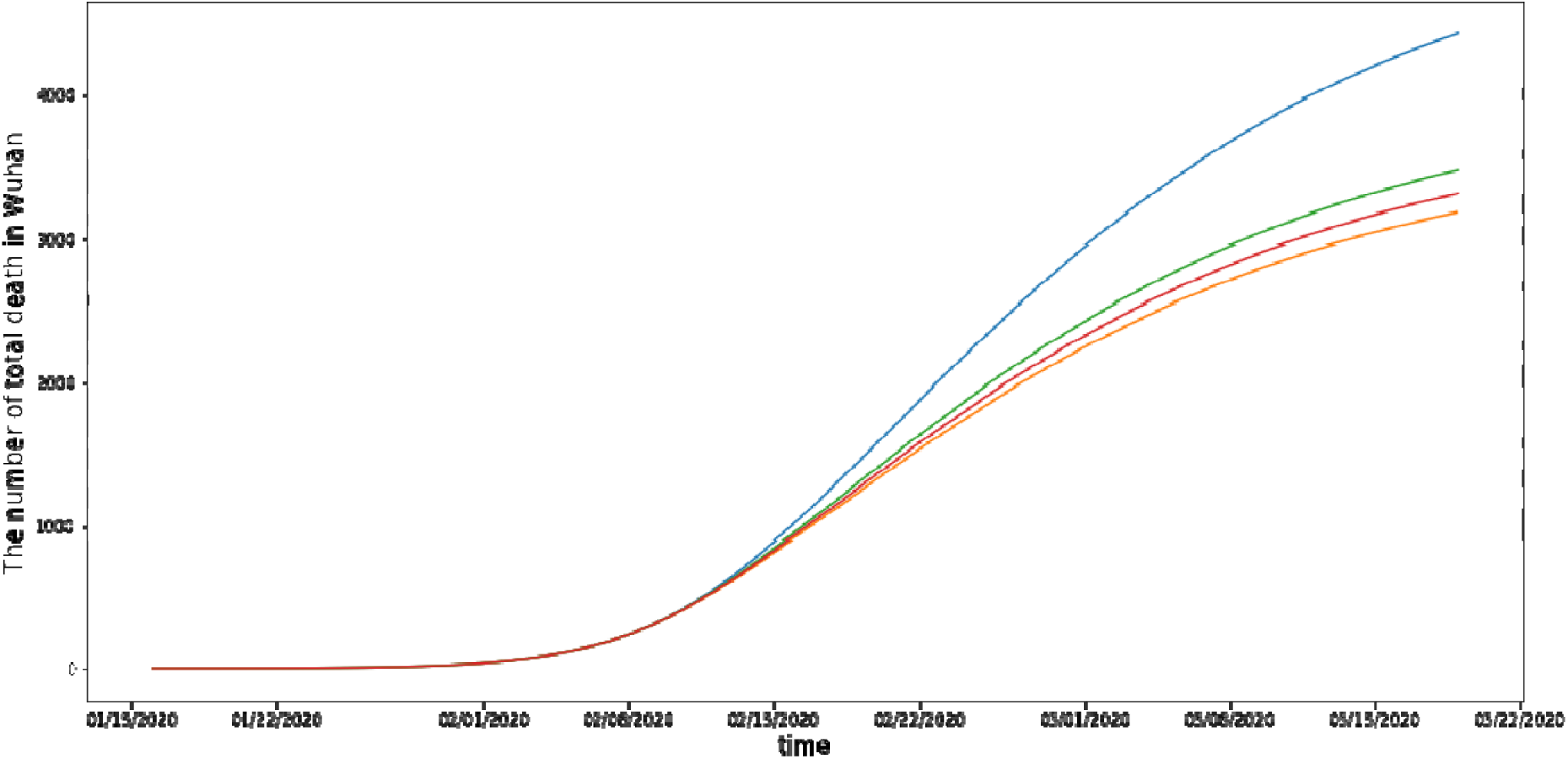
Sensitivity analysis of the effect of capacity on the number of deaths in Wuhan. Note: The blue, green, red, and orange lines denote the predicted number of deaths in Wuhan with the original hospitalized rate, a 50% increase in the hospitalized rate, a 75% increase in the hospitalized rate, and a 100% increase the hospitalized rate, respectively.

## 5. Additional Analyses in New York State and Italy

With the outbreak of COVID-19 epidemics in the North America and Europe, medical capability shortages were reported in New York State in the United States and in Italy. We used the situation at these two locations to demonstrate the generalizability of the model with capacity limits at the state and country levels.

### 5.1 Data and Parameter Setting in New York State and Italy

To estimate the situations, we collected the numbers of infected and hospitalized patients in New York State between March 8 and April 27, 2020, from the US Centers for Disease Control and Prevention^3^ and those in Italy between February 23 and April 24, 2020, from the country’s Civil Protection Department.^4^ Because of the lack of individual recovery records in the New York state, we used the reported average recovery period, namely 12 days (Guan et.al, 2020).

We also collected their tested numbers to validate our starting point, turning points and ending points for the change of beta and fraction (i.e., *t*_0_,*t*_1_, *t*_2_,*T*_0_,and *T*_1_). We set *t*_0_, *t*_1_,*t*_2_, *T*_0_,and *T*_1_ at 72, 105, 72, 105, and 135, respectively, for New York State and 60, 100, 60, 100, and 135 for Italy. In addition, because a home-isolation policy was executed in New York State and Italy and only patients with severe symptoms would be hospitalized, the final hospitalized rate did not increase to one but fluctuated below one. Thus, we set the parameter *U* in eq. (1) at 0.65 for New York State and Italy according to the trend for the hospitalized rate over the number of infected patients. Furthermore, in the SEIR model, we set the parameters *γ* and *θ* to 0.08 and 0.25, respectively, which were derived from the model using Wuhan data. We also set *N* in New York State and Italy at the respective population numbers.

### 5.2 Estimated Results in New York State and Italy

The *R*^2^ values of the models for New York State and Italy were 0.996 and 0.972 respectively, indicating a good fit within the data set. The fitted *d*_1_ and *d*_2_ were 0.075 and 0.349 for New York State and 0.040 and 0.375 for Italy, respectively, which are close to the fitted parameters for Wuhan. The higher *d*_2_ for Italy indicates that it improved its capacity more quickly than other regions did.

Fig. 9 and Fig. 10 illustrate the changes of transmission rate and hospitalized rate, respectively, in New York State and Italy. The results revealed that the *β*_0_ in New York was 0.506 at *t*_0_ = 72, which is higher than the initial transmission rate of 0.334 in Italy at *t*_0_ = 60. This difference suggests more frequent contact between people in New York State before control measures were introduced.

**Fig. 9.**
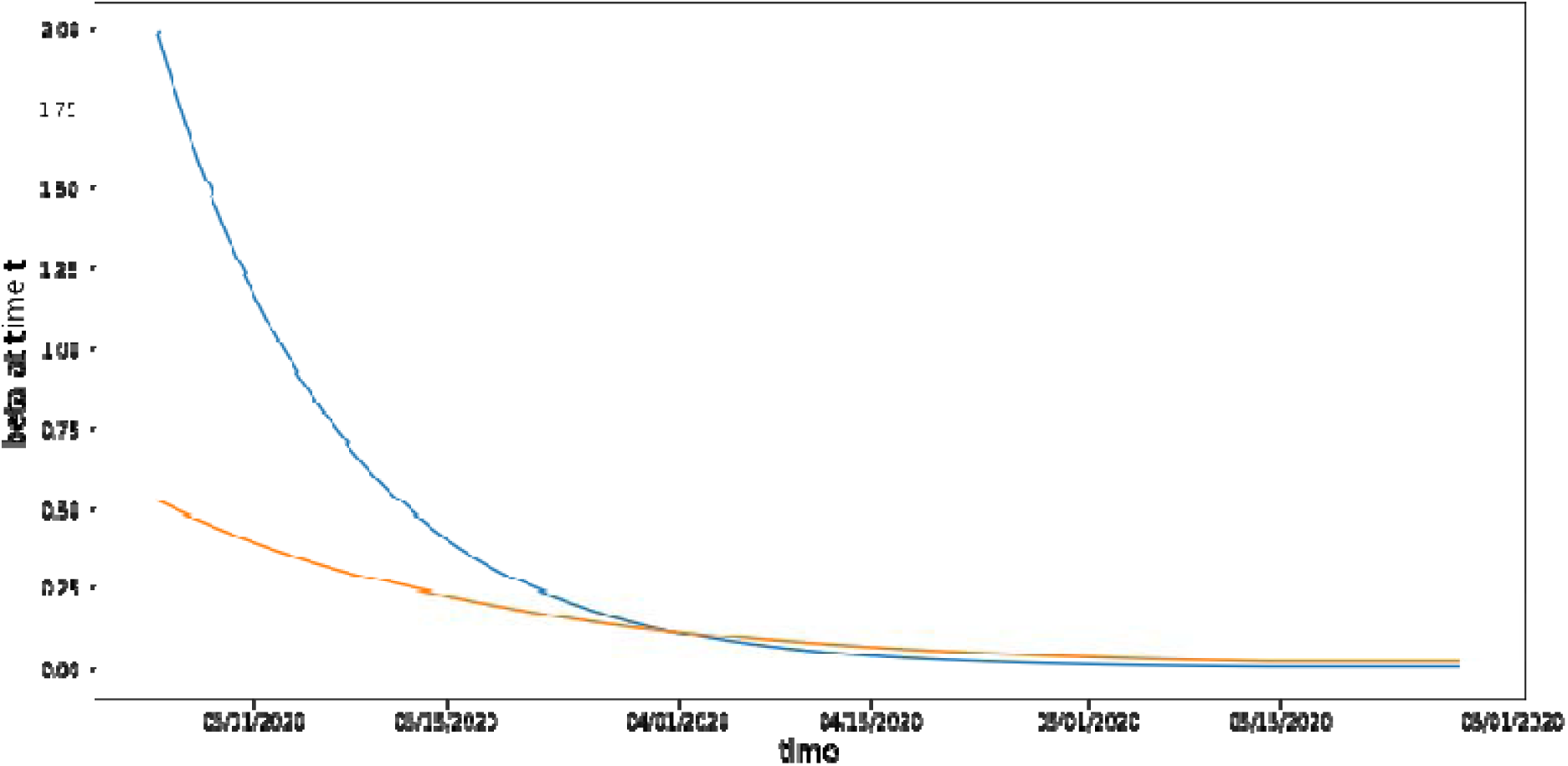
Transmission rates in New York State and Italy. Note: The blue and orange lines denote the transmission rates in New York and Italy, respectively.

**Fig. 10.**
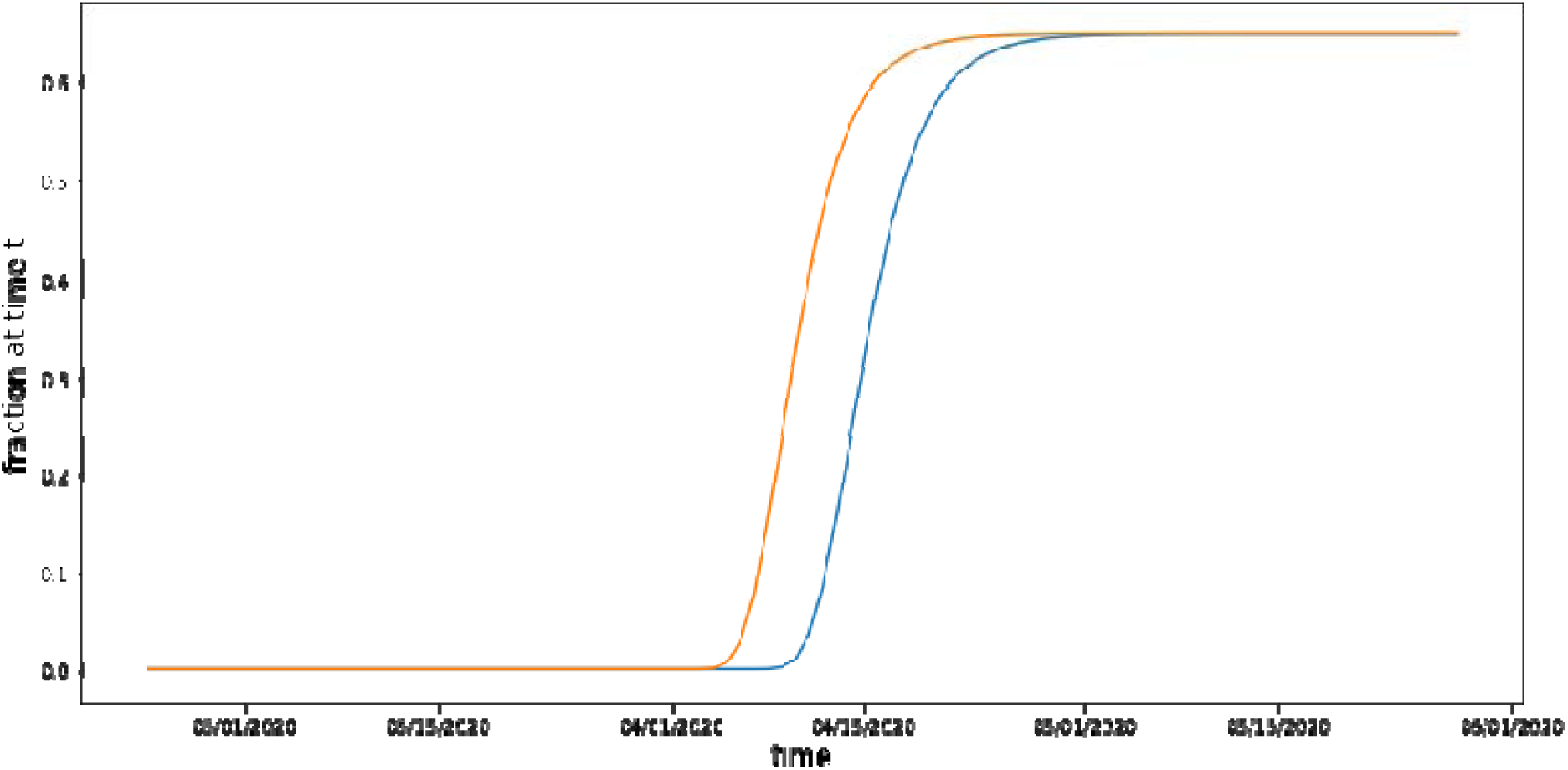
Hospitalized rate in New York State and Italy. Note: The blue and orange lines denote the hospitalized rates in New York State and Italy, respectively.

The average transmission rate in New York State before April 2020 was approximately 2.5 times that in Italy. Furthermore, the difference in transmission rates meant that New York State reached the 0.20 transmission rate of *β* 7 days later than Italy did. Moreover, Fig. 10 indicates that the process of improvement in medical capacity in New York State had a 6-day delay in reaching a 60% hospitalization rate compared with Italy. For New York State, the hospitalized rate increased from 1.15% on April 11 to 64.5% on April 29, 2020, whereas for Italy, the hospitalized rate increased from 2.98% on April 7 to 64.5% on April 23, 2020.

We also conducted sensitivity analyses in the cases of New York State and Italy. Specifically, we varied the speed at which the transmission rate decreased by multiplying the original *d*_1_ by a ratio ranging between 0.75 and 1.5. The results, as illustrated in Fig. 11(a), indicate that a 25% slower decrease in the original transmission rate in New York State would lead to a 238.88% increase in the peak number of infected patients, which is much higher than that in Wuhan. In addition, a 25% or 50% faster decrease in the original transmission rate would lead to 52.08% or 69.46% smaller peak number of infected patients, respectively. As shown in Fig. 11(b), In Italy, a 25% slower decrease in the original transmission rate would lead to a 228.32% increase in the peak number of infected patients, whereas a 25% or 50% faster decrease in the transmission rate would lead to a 57.73% or 76.11% decrease in the peak number of infected patients, respectively. A possible reason for why slowed transmission rates had a greater influence in New York State and Italy than it did in Wuhan is the higher starting transmission rates and longer time before the decline in transmission rate (*β*) in New York State and Italy.

**Fig. 11.**
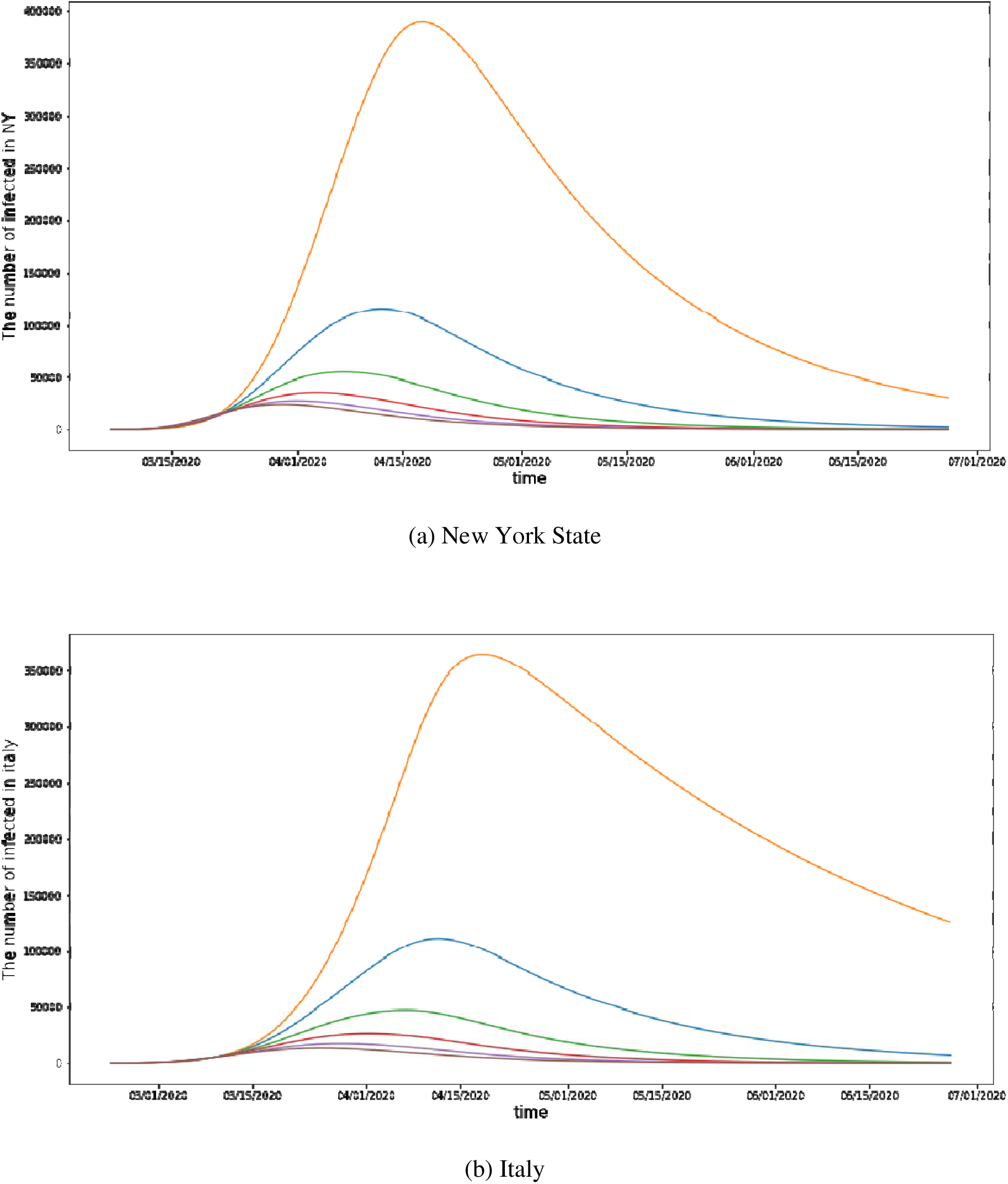
Model estimation with changing transmission rate in New York State and Italy. Note: The blue, orange, green, red, purple, and brown lines denote the number of infected patients with the original, a 25% lower, a 25% higher, a 50% higher, a 75% higher, and a 100% higher, respectively.

For the sensitivity analysis on medical capacity, we varied the hospitalized rate to 0.25, 0.5, 0.75, 1.5, 1.75, and 2 times of their original levels, respectively, and Fig. 12 presents the results. Because the change in rate would influence the time point of the peak number of infected patients as well as the rate at which the infection rate slows, we evaluated not only the peak number of infected patients but also the speed at which it decreased. Regarding the peak in New York State, 75%, 50%, and 25% decreases in the hospitalized rate would lead to increases in the peak number of infected patients from 11,497 to 115,049, 115025, and 115,001, respectively. Moreover, 50%, 75%, and 100% increases in the rate would reduce the peak number to 114,929, 114,905, and 114,881 respectively. For Italy, 75%, 50%, and 25% decreases in the hospitalized rate would lead to increases in the peak number from 110,982 to 117,108, 113,578, and 115,001, respectively. Moreover, 50%, 75%, and 100% increases in the hospitalized rate would reduce the peak number of infected patients to 109,649, 109,121, and 108,774, respectively.

**Fig. 12.**
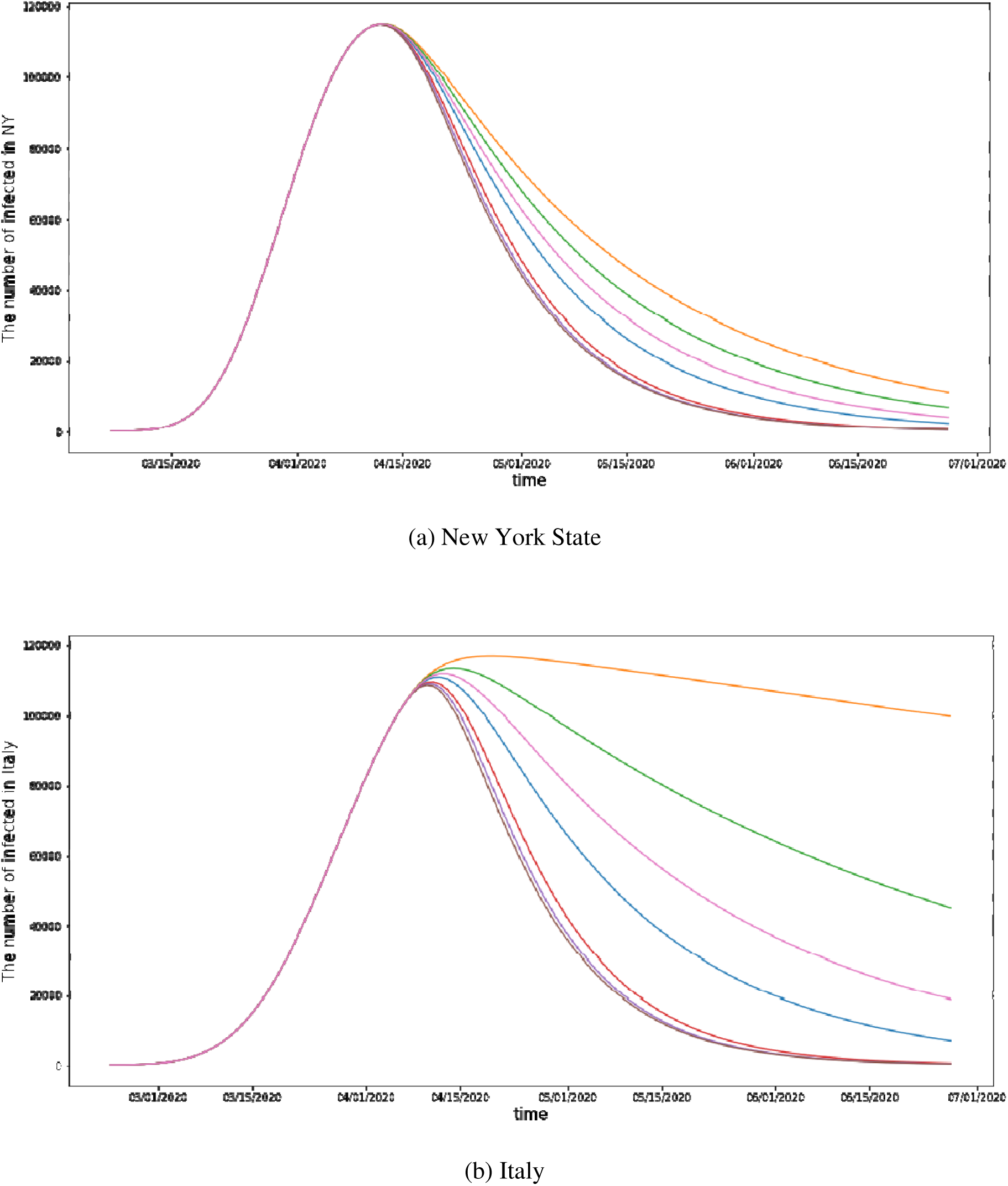
Sensitivity of hospitalized rate in New York State and Italy. Note: The blue line denotes the number of infected patients with the original hospitalized rate. The orange, green, and pink lines denote the number of infected patients with 75%, 50%, and 25% lower hospitalized rate, respectively. The red, purple, and brown lines denote the number of infected patients with 50%, 75%, and 100% higher hospitalized rate, respectively.

To determine the effects of different hospitalized rates on the speed of decline in the infection rate, we also examined the time at which the number of infected patients would decrease to half of the peak value. The time point at which the peak number in New York State would decrease to below 57,500 (i.e., approximately half of its estimated peak number) would be May 3 based on the original rate, and it would be delayed to May 10, May 7, and May 4 for 75%, 50%, and 25% decreases in the hospitalized rate, respectively. Increasing the hospitalized rate in New York State by 50%, 75%, and 100% would move the time point for reaching 50% of the peak forward to April 30, April 29, and April 29, respectively. For Italy, the original time point for the number of infected patients to decrease to below 55,500 (i.e., by approximately half of the peak number) would be May 7. This would be delayed to July 28, July 14, and May 17 if the hospitalized rate were 75%, 50%, and 25% smaller respectively. Increasing the hospitalized rate by 50%, 75%, and 100% would cause the half-peak level to arrive on April 28, April 27, and April 26, respectively.

Furthermore, changes in the hospitalized rate would influence the total number of deaths in New York State and Italy. From the concluded cumulative total numbers of positive cases and deaths, we were able to estimate the death rate to be 0.084 in New York State and 0.135 in Italy. Accordingly, we found that 75%, 50%, and 25% decreases in the hospitalized rate in New York State would lead to increases in the number of deaths from 28,817 to 35,785, 33,003, and 30,712, respectively (see Fig. 13). Furthermore, 50%, 75%, and 100% increases in the hospitalized rate would lead to decreases in the number of deaths to 25,919, 25,346, and 25,088, respectively. Moreover, 75%, 50%, and 25% decreases in the rate in Italy would lead to increases in the number of deaths from 34,913, to 69,223, 52,799, and 42,034, respectively; furthermore, 50%, 75%, and 100% increases in the hospitalized rate would lead to decreases in the number of deaths to 26,783, 25,635, and 25,179, respectively.

**Fig. 13.**
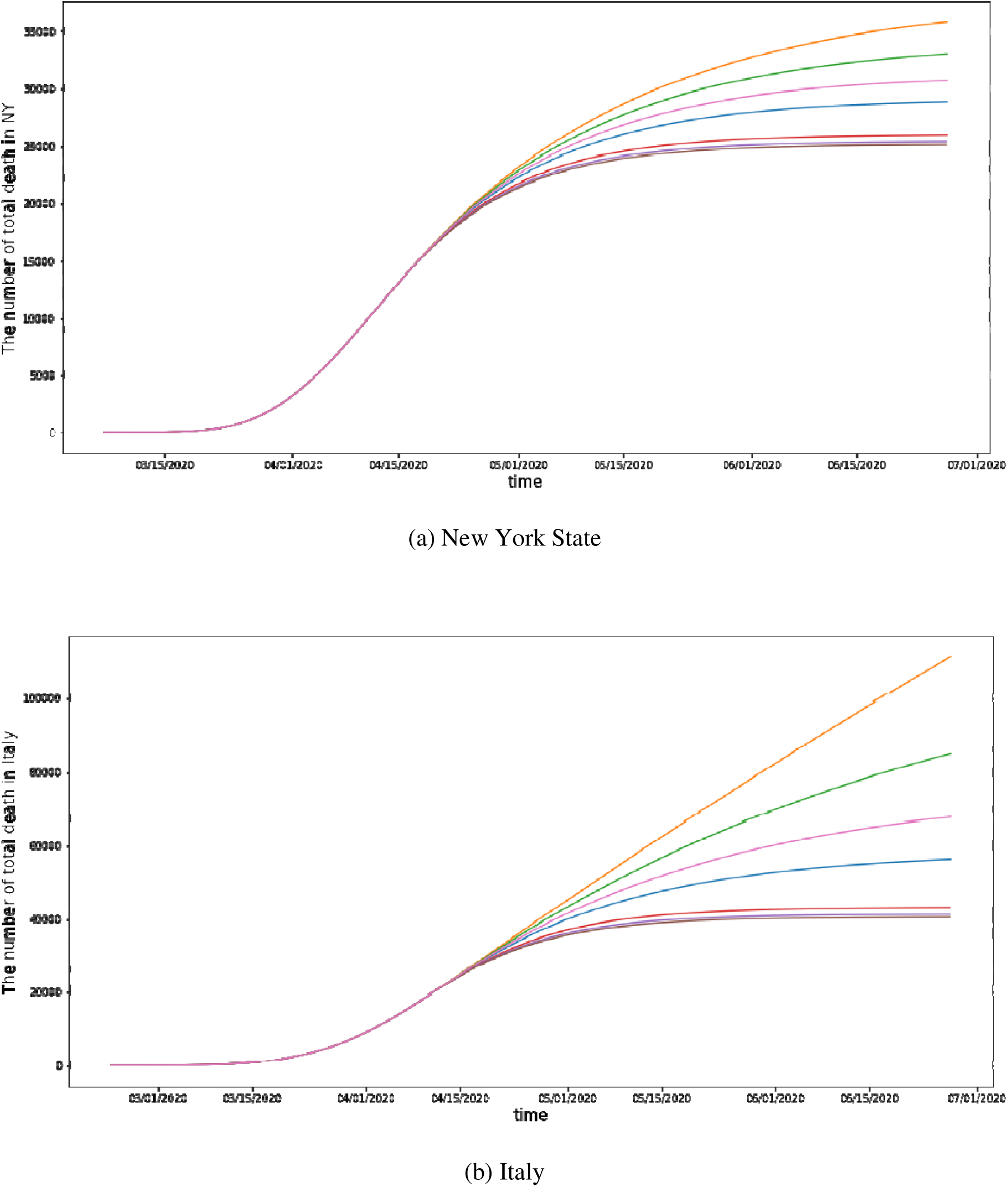
Model estimation of total deaths with changing rates in New York State and Italy. Note: The blue line denotes the number of deaths with the original hospitalized rate. The orange, green, and pink lines denote the number of deaths with 75%, 50%, and 25% decreases in the hospitalized rate, respectively. The red, purple, and brown lines denote the number of deaths with 50%, 75%, and 100% increases in the hospitalized rate, respectively.

## 6. Robustness Checks

We conducted three robustness checks in this study. The first one tested the influences of different values of U on the epidemics in New York State and Italy, the second robustness check accounted for the influence of human mobility from central infection region to outside regions, and the last one considered different solvers used in the process of parameter searching.

First, as defined earlier, *U* is the maximum hospitalized rate as a percentage of all confirmed infections and it can vary in the range between zero and one. In the cases of New York State and Italy, we set the value of *U* to be 0.65 in the main analysis since the governments there implemented home-isolation policies in which only patients with confirmed infections and severe symptoms were hospitalized while other patients underwent home isolation. In other regions such as China, the value of *U* was set as one, which represents the situation of all confirmed infections being hospitalized, with no infected patients undergoing home isolation. In this robustness analysis, we varied the value of *U* at a set of different levels: 0.25, 0.5, 0.75, and 1 respectively. The corresponding results are presented in Fig. 14.

**Fig. 14.**
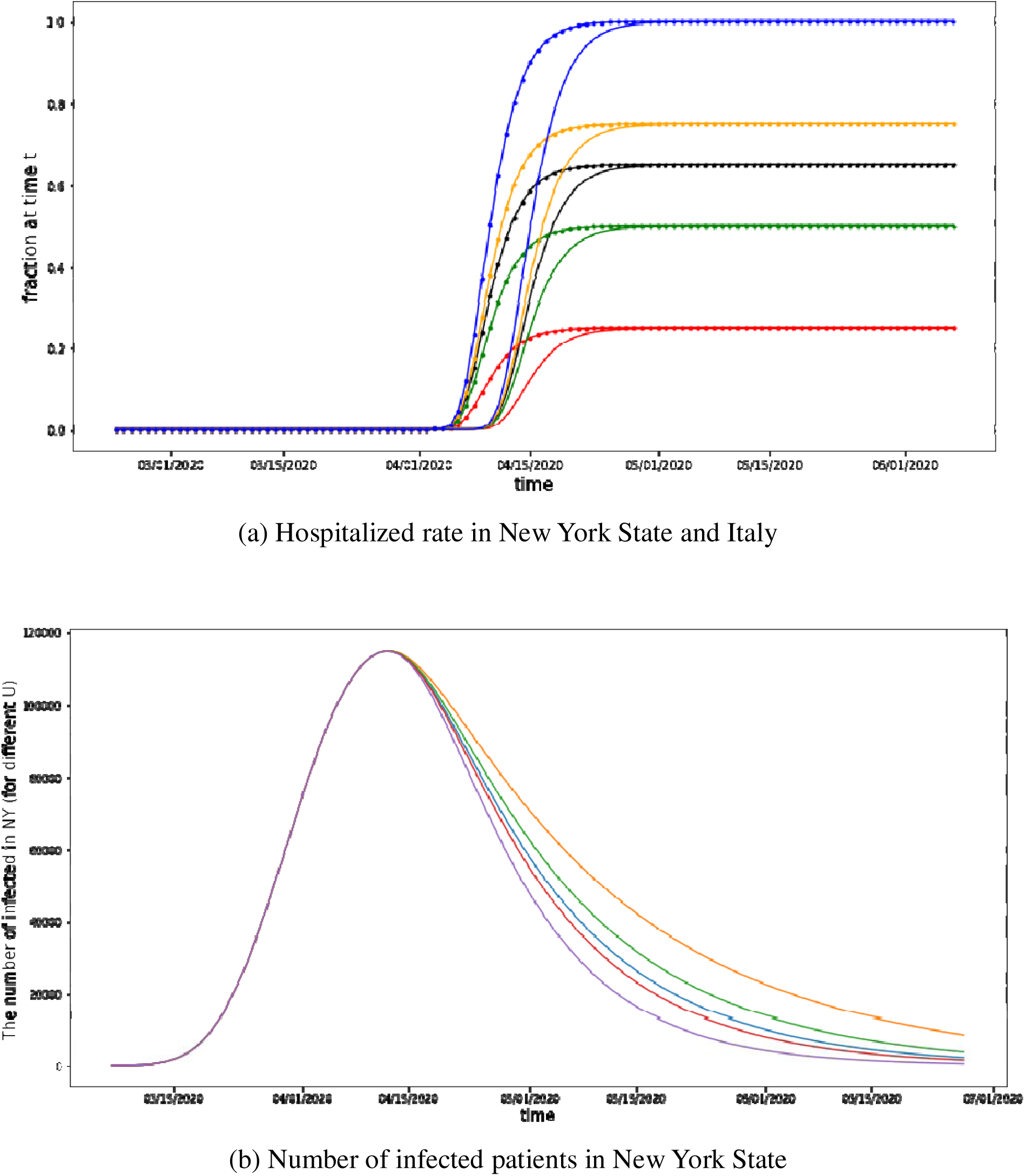
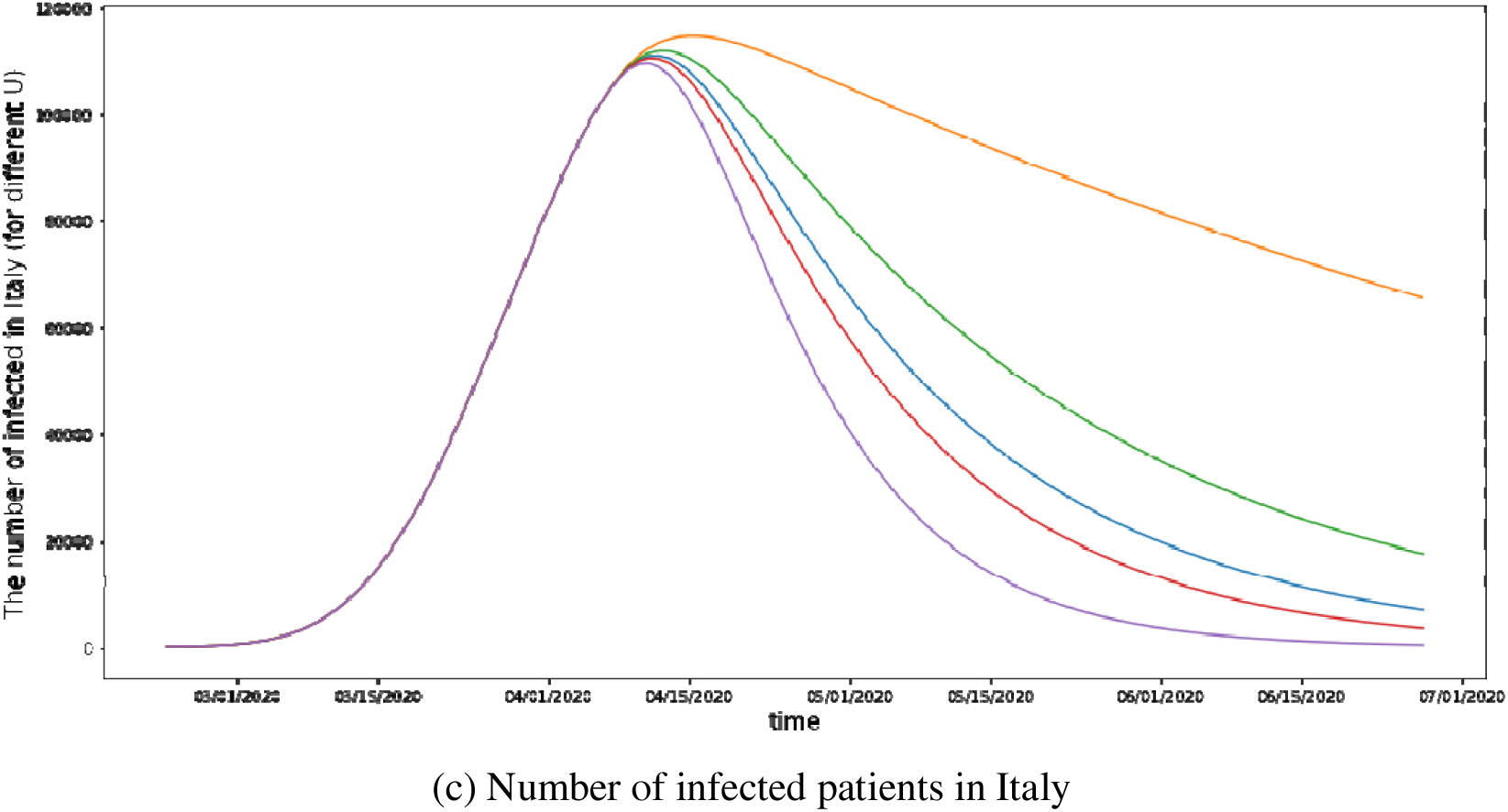
Effects of different U values on infection numbers in New York State and Italy. Note: In (a), the black, red, green, orange, and blue lines denote the hospitalized rate for the original *U* (0.65), *U* = 0.25, *U* = 0.5, *U* = 0.75, and *U* = 1 respectively. The dotted line denotes Italy and the ordinary line denotes New York State. In (b) and (c), the blue, orange, green, red, and purple lines denote the number of infected patients with the original *U* (0.65), *U* = 0.25, *U* = 0.5, *U* = 0.75, and *U* = 1 (the ideal case). respectively.

As shown in Fig. 14(a), changes in *U* would result in a proportional change in the hospitalized rate. It would also influence the epidemic curve for both New York State and Italy; see Fig. 14(b) and Fig.14(c) respectively. For New York State, reducing *U* to 0.25 and 0.5 would increase the peak number of infected patients from 114,977 to 115,036 and 114,999, respectively. Moreover, increasing *U* to 0.75 and 1 would increase the peak number to 114,963 and 114,926, respectively. For Italy, reducing *U* to 0.25 and 0.5 would increase the peak number of infected patients from 110,982 to 114,708 and 111,886, respectively. Moreover, increasing *U* to 0.75 and 1 would reduce the peak number to 110,500 and 109,558, respectively.

In addition, changes in *U* would also influence the total number of deaths. Fig. 15 indicates that reducing *U* to 0.25 and 0.5 in New York State would lead to increases in the number of deaths from 28,817 to 34,220 and 30,553, respectively; increases in *U* to 0.75 and 1 would lead to reductions in the number of deaths to 27,813 and 25,735, respectively. Moreover, reducing *U* to 0.25 and 0.5 in Italy would lead to increases in the number of deaths from 34,913 to 59,513 and 41,378, respectively; increasing *U* to 0.75 and 1 would lead to reductions in the number of deaths to 31,742 and 26,364, respectively.

**Fig. 15.**
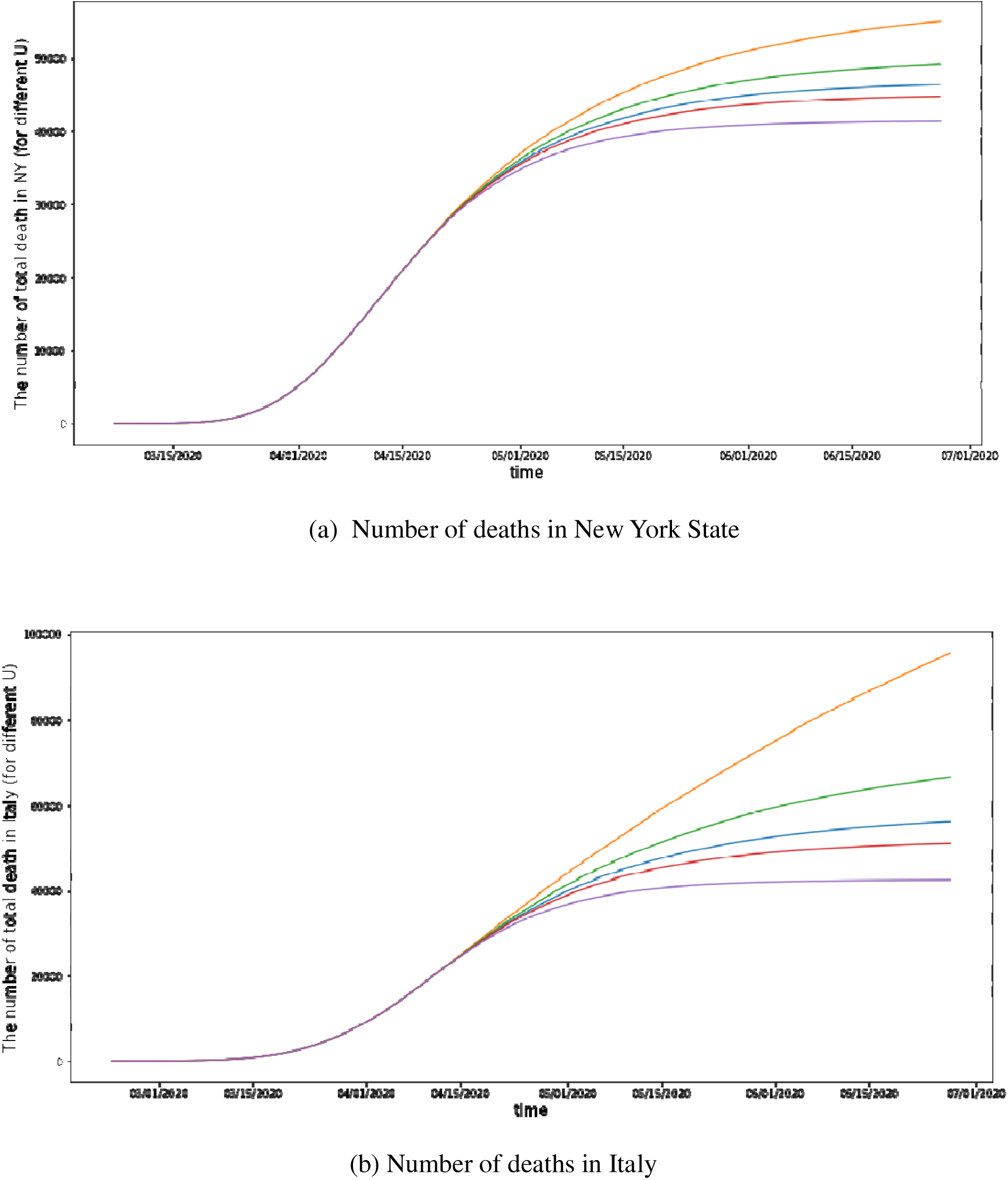
Effects of different U values on the number of deaths in New York State and Italy. Note: The first graph represents New York State, and the second graph represents Italy. The blue, orange, green, red, and purple lines denote the number of deaths with the original *U* (0.65), *U* = 0.25, *U* = 0.5, *U* = 0.75, and *U* = 1, respectively.

Our second robustness check further considered whether human mobility would affect the model’s validity. Specifically, we extended the proposed dynamic SEIR model by introducing the spatial spread of the virus across regions on the basis of human mobility data from the central epidemic area to other cities. This is because that regions outside the central area (such as regions of China outside of Hubei Province) were affected by cases exported from the center (such as Wuhan) [3, 4], and such human mobility would increase the exposure rate in cities outside of the central area. Thus, eq. (4) can be rewritten as follows:

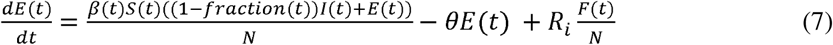

where *F*(*t*) and *R*_*i*_ denote the human travel flow from the central area to outside region *i* at time *t* and the corresponding rate of infected people traveling from the central area to other region *i*, respectively. This equation especially applies to cases in which people travel from a highly infected region to a noninfected region (such as during the 2020 Spring Festival travel season). Without losing generality, the capacity shortage time *t*_1_ and measure action time *t*_2_ would differ (usually several days later) from those in the central area. Furthermore, we assumed that the number of infected people traveling from the central area to other regions would vary across regions and thus that the transmission rate would be specific to the destination region *i*.

We applied the same parameter settings as those adopted in the main analysis. Human mobility data were collected from the Baidu Migration Index in 2020, which measures travel flow through changes in mobile phone locations. The performance of the new model in terms of accuracy and explanatory power was largely consistent with that displayed in Fig. 4. Therefore, accounting for mobility did not affect the main findings.

In the last robustness check, we compared Nelder–Mead method, which is used in our analysis, with Conjugate Gradient (CG) method, Broyden–Fletcher–Goldfarb–Shanno (BFGS) algorithm, and Limited-memory BFGS (L-BFGS). CG is a modified gradient descent method based on conjugate directions of gradient descent. It avoids the computation of Hessian Matrix and its inverse, which leads to higher efficiency and robustness. BFGS is a type of Quasi-Newton method, which avoids the computation of Hessian through approximation. L-BFGS limits history size of iteration compared with BFGS, and it uses a relatively sparse implicit representation of Hessian approximation. All these methods are widely used in nonlinear optimization problems. Table 1 summarizes the R-Squared values when using these methods in our analysis, which demonstrate that the Nelder–Mead method performs significantly better than other three methods.

**Table 1:** Comparison of R-squared values when using different fitting methods.

| **Methods** | ***Study Regions*** |  |  |  |
| --- | --- | --- | --- | --- |
|  | **Wuhan** | **Outside of Hubei Province** | **New York State** | **Italy** |
| **Nelder-Mead** | 0.984 | 0.926 | 0.996 | 0.984 |
| **CG** | 0.766 | Not solved | 0.893 | 0.516 |
| **BFGS** | 0.851 | Not solved | 0.893 | 0.525 |
| **L-BFGS** | 0.799 | Not solved | 0.727 | 0.582 |

The major difference between these methods is that the Nelder–Mead method is a direct search method that does not need to calculate derivatives, and thus its performance on non-linear functions is better than derivative-based methods, especially when the derivatives are hard to compute. In our model, the Gompertz functions in the fraction and β equations have brought in some type of non-linearity and made it more difficult to calculate the derivatives, and thus it is natural that the Nelder–Mead method performs better in our problem.

## 7. Discussion

Medical capacity shortages are a considerable challenge faced by many public health systems during the COVID-19 pandemic and other epidemics that will likely arise again in the future. Through the development of a novel SEIR-based model, this study investigated the impact of medical capacity shortages and empirically estimated their severity in Wuhan, New York State, and Italy during the outbreak and first wave of COVID-19 pandemic. Among these locations, Italy faced the most severe medical capacity shortages given its lower hospitalization fraction in the rising phase of the pandemic.

Using observable infections and medical capacity information, this study demonstrates that the shortage in medical capacity significantly increased the infected number; with Wuhan as the example, the infection number would be 39% lower than the actual number if the capacity were twice of the original size. This is the first quantified evidence to show that bed shortages facilitate the spread of COVID-19. To reduce the impact of this type of shortage, public health systems should be flexible in increasing their medical capacity. Increased recruitment of retired doctors and nurses and the establishment of mobile field hospitals [17-19] have been implemented to compensate for deficiencies in medical capacity and slow the spread of COVID-19. We believe it is still critical to manage the medical capacity in the coming second wave of COVID-19 pandemic.

If the shortage cannot be resolved promptly and efficiently, the infection curve of the pandemic will inevitably steepen. While affected countries are making efforts to produce or import medical equipment, chronic global shortages of various medical equipment and medical staff remain one of the most urgent threats to numerous countries. Uniting to confront the pandemic is necessary. Instead of fighting alone, different countries and regions should combine their efforts through a more coordinated supply chain and resource sharing with international or interregional cooperation.

Our robustness check on *U* revealed the importance of medical system and patient treatment strategies during the pandemic. A small *U* could represent a lax attitude to COVID-19, whereas a large *U* may indicate strict control measures and aggressive isolation policies, such as staying-home notices, acceptance of and provision of treatment for all patients, and wide-range swab testing. Compared with a larger *U*, a small *U* would lead to a consistent pandemic. Taking Italy as an example, if *U* = 0.75, the total patient number (I + R + E) would equal 381,978 on July 1, whereas this number would be more than double if *U* = 0.25.

Crucially, other measures indicate that the situation is changing daily. The findings of this study also reveal how control and prevention measures may moderate the COVID-19 spread. While various pharmaceutical and non-pharmaceutical interventions such as social distancing measures, health practice promotion, and supply chain enhancement have been implemented to reduce the transmission rate, medical capacity shortages could limit their positive effects.

Many characteristics of COVID-19 remain unknown or uncertain. Consequently, care should be taken when interpreting our estimates. For instance, various factors affect medical capacity. In addition to the number of general beds, the number of intensive care beds is thought to be more critical for severe cases. Moreover, personal protective equipment and ventilators might be other types of medical resources affected by shortages during different periods of the COVID-19 pandemic. Further studies should account for more factors associated with medical capacity.

## Data Availability

The data will be posted on the online repository shortly

## Footnotes

1 Data were collected from the daily information announced by provincial health commissions in China during the COVID-2019 pandemic. The data of bed numbers in the whole country were obtained from City Statistics Yearbook.

2 The calculation of average treated fraction: use arithmetic average(*f*_1_ + · + *f*_*k*_)/*k*, where *f*_*i*_ is the treated fraction at day *i*, and *k* is the total number of days we take into consideration.

3 The data were collected from https://covidtracking.com/api

4 The data were collected from https://github.com/pcm-dpc/COVID-19

